# Muscle bursting and corticomotor excitability index impaired impulse control in Parkinson’s disease

**DOI:** 10.1101/2025.04.28.25326550

**Authors:** Aliya C. M. Warden, Craig McAllister, Damian Cruse, Ben Wright, Hayley J. MacDonald

## Abstract

Dopamine agonist medications in Parkinson’s disease pose a significant risk for the development of impulse control disorders. However, the neural mechanisms underlying impaired impulse control while on agonist medication remain unclear, limiting our ability to pinpoint measures which predict impulse control disorder onset. This study aimed to identify objective markers of impaired impulse control in people with Parkinson’s by examining corticomotor excitability and muscle bursting during an impulse control task.

Nineteen people with Parkinson’s on ropinirole (64.1 ± 7.0 years, 8 female, Hoehn and Yahr stages 1–2.5) and 18 healthy older adults (69.1 ± 5.6 years, 9 female) completed the anticipatory response inhibition task in this cross-sectional study, requiring response withholding (Go trials) and inhibition (Stop trials). Transcranial magnetic stimulation was applied during both trial types, while corticomotor excitability and muscle bursts, recorded via electromyography, were measured during response withholding and successful stopping.

During response withholding, people with Parkinson’s exhibited an early and transient increase in corticomotor excitability alongside a higher rate of dysfunctional muscle bursts compared to controls, which led to more variable lift responses. During response inhibition, people with Parkinson’s demonstrated a smaller reduction in corticomotor excitability and a greater frequency of maladaptive muscle bursts following the stop-signal, which led to delayed inhibition of the response. Greater impairments in response inhibition were associated with a higher frequency of impulse control disorder behaviours.

The current study identified neurophysiological measures sensitive to deficits in response withholding and inhibition in Parkinson’s disease. Collectively, impaired task performance was characterised by reduced suppression of corticomotor excitability and increased dysfunctional muscle bursting. These changes were associated with greater disease severity and higher dopaminergic medication doses and thus may reflect overdosing effects on the inhibitory control network. Our findings highlight the potential of these neurophysiological measures to serve as early objective markers of impulse control dysfunction in Parkinson’s disease. Further investigation into the predictive capability of these markers for the development of impulse control disorders will help establish their use as a valuable tool in clinical practice.

## Introduction

Parkinson’s disease (PD) is a neurodegenerative disorder affecting 1–2 per 1000 people, with incidence rising sharply after 65.^1,2^ While PD is characterised by motor symptoms such as bradykinesia, tremor, and rigidity, non-motor symptoms are also common, including impulse control disorders (ICDs). Dopamine agonists (DAs), especially ropinirole and pramipexole, are effective for motor symptoms but strongly associated with ICDs^3,4^ (∼42% incidence)^5^, likely via overstimulation of D_3_ receptor-mediated reward pathways.^6^ These ICD behaviours—pathological gambling, binge eating, compulsive shopping, and hyper-sexuality—can severely impact quality of life, relationships, and financial stability.^7,8^ Currently, no objective method exists to identify those at risk.

Impulsivity encompasses two main domains: impulsive choice (impaired decision-making) and impulsive action (inability to suppress prepotent responses)^9^. Impulsive action is typically assessed through response inhibition paradigms such as the Go/No-Go and Stop-Signal tasks, which require withholding and cancellation of externally cued responses, respectively. PD-related impairments have been documented across both tasks,^10–15^ including under DA treatment and before ICDs manifest,^9,16^ although there are some exceptions.^14,17^ People with PD (PwPD) have particular difficulty with internally generated, anticipatory responses, likely due to reduced preparatory suppression—corticospinal inhibition that typically prevents premature response initiation.^18,19^

Control of such anticipated responses can be examined using the Anticipatory Response Inhibition Task (ARIT) which is becoming increasingly popular.^20^ In Go trials, response withholding must be proactively maintained and then strategically released to initiate movement synchronising with a stationary target. In contrast, Stop trials require reactive inhibition of the prepared response, triggered by the presentation of the stop-signal. Therefore, the ARIT provides insight into both proactive and reactive aspects of inhibitory control, which interact dynamically to enable adaptive behavioural control.^21^ To our knowledge, only one study has applied the ARIT in PD,^22^ demonstrating that poorer response inhibition performance was associated with a higher frequency of ICD behaviours on DA medication.

The neurophysiological mechanisms underlying impaired performance in the ARIT for PwPD remain unclear. Investigating these mechanisms could help to identify objective markers of impaired impulse control associated with everyday impulsive behaviour. For instance, electromyography recordings during the ARIT suggest that PwPD are more likely than age-matched controls to exhibit ineffective bursts of muscle activity when withholding the planned response in Go trials (see^23^). It is possible that this reflects dysfunction within the inhibitory control network, specifically preparatory suppression, and warrants further investigation in a full cohort.

Bursts of muscle activity during Stop trials also provide insight into underlying neurophysiological mechanisms; in this case during the reactive stopping process. In successful Stop trials, no behavioural response occurs, as the prepotent movement is successfully inhibited. However, participants may still generate sub-threshold muscle activation, initiating but subsequently cancelling the motor response before sufficient force is generated to carry out the action. The latency of these bursts relative to the stop-signal (CancelTime) directly indexes action stopping at a single-trial level. These partial muscle bursts occur at rates of ∼30-35% in healthy adults performing the ARIT,^24,25^ and can be observed more frequently in clinical groups with basal ganglia dysfunction e.g., focal dystonia.^26^ Overall, we propose that muscle bursting during both response withholding and inhibition may serve as a physiological marker of inhibitory control in PwPD.

To better understand muscle-bursting activity in PD, it is essential to explore further upstream in the motor control pathway. Many studies have applied single-pulse transcranial magnetic stimulation (TMS) over the motor cortex (M1) to record changes in corticomotor excitability (CME) during impulse control tasks in healthy adults (see^27^ for a review).^28,29^ Specifically in the ARIT, following a stable baseline period, CME increases around 250-175 ms before the target and remains elevated in Go trials.^30–32^ In contrast, successful response inhibition is associated with a reduction in CME 140-175 ms following stop-signal presentation.^31,33^ To our knowledge, no studies have investigated how CME fluctuations in PwPD compare to the typical pattern observed during response inhibition tasks.

Overall, the neural mechanisms underlying impaired impulse control in PwPD remain unclear, despite a high prevalence of destructive ICDs in this population. Therefore, the present study aimed to identify objective markers of impaired impulse control in PD by examining changes in muscle bursting and CME measures during response withholding and inhibition. A further aim was to investigate whether these objective markers were associated with everyday impulsive behaviour. Our hypotheses were as follows; compared to healthy older adults, PwPD on the DA ropinirole would show: (1) an impaired ability to a) withhold and b) inhibit the prepared lift response in the ARIT; (2) more ineffective muscle bursting activity during a) response withholding and b) response inhibition; (3) a) increased CME during response withholding and b) reduced CME suppression during response inhibition; and (4) a link between muscle bursting and CME changes with task performance and self-reported impulsivity.

## Materials and methods

### Participants

Twenty healthy controls (HCs) and 23 PwPD on ropinirole were recruited as part of a larger pre-registered study (see https://doi.org/10.17605/OSF.IO/KC2H3 and https://doi.org/10.17605/OSF.IO/W2FHP and Supplementary Material 1 for more details). One PwPD was excluded due to cognitive impairment, while three PwPD and two HCs were excluded due to outlying behavioural values or unreliable neurophysiological measures. Therefore, 18 HCs (69.1 ± 5.6 years, 9 female, 16 self-reported as right-handed) and 19 individuals (64.1 ± 7.0 years, 8 female, 16 self-reported as right-handed) with a clinical diagnosis of PD (Hoehn and Yahr stages 1–2.5) contributed data to the study. A minimum group size of 15 was determined via power analysis (α = 0.05, 80% power) based on a medium effect size (Cohen’s *d* = 0.69) reported in prior CME research.^31^

HCs were recruited via the Birmingham 1000 Elders group and PwPD via the Parkinson’s UK research network and routine NHS clinic appointments across England, Wales and Scotland. Participants were recruited if aged 40-80, with no history of neurological conditions other than PD or reported vision impairment that was not corrected (e.g., with glasses). Participants were screened for TMS eligibility based on current international safety guidelines^34^ and required a score >23 on the Montreal Cognitive Assessment (MoCA).^35^ No PwPD had received deep brain stimulation. The Movement Disorder Society-Sponsored Revision of the Unified Parkinson’s Disease Rating Scale (MDS-UPDRS; parts I-III),^36^ assessed symptom severity while PwPD were on their usual medication. The Hoehn and Yahr scale assessed functional disability.^37^

All participants gave written informed consent and received monetary compensation for their participation. The study received favourable opinion from an NHS Research Ethics Committee (South-East Scotland REC 02, IRAS ID: 328075) and was conducted in accordance with the Declaration of Helsinki.

### Behavioural task

The current study used the bimanual version of the ARIT (Fig. 1), controlled via custom MATLAB software (R2019b, The MathWorks) and interfaced with two custom-made microswitches. The medial aspect of each index finger was used to lightly depress the two switches (index finger adduction), with the left and right indicators corresponding to the respective fingers. After a 2 s delay, both indicators started to move upward from the bottom at equal rates.

**Figure 1.**
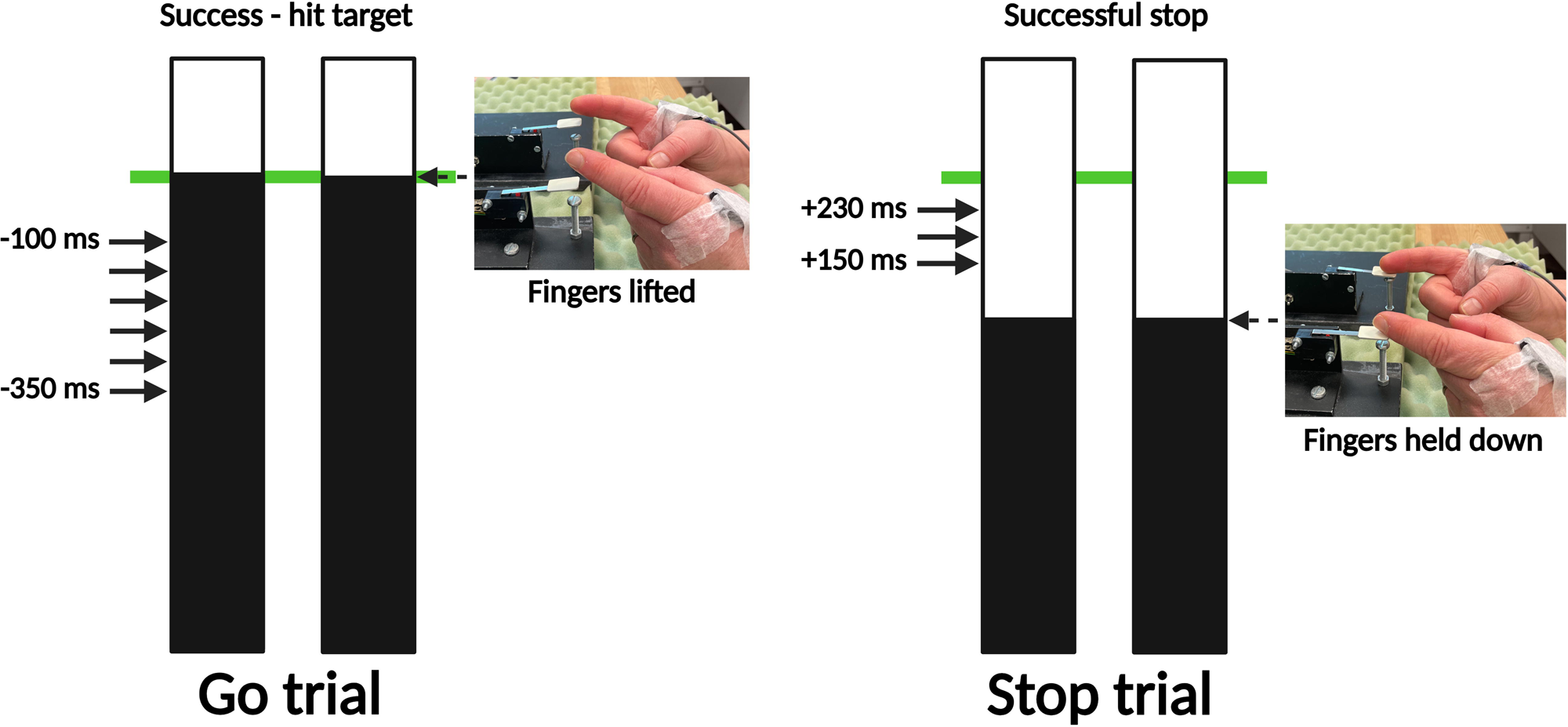
The Anticipatory Response Inhibition Task. During Go trials, participants initially held their index fingers on the respective switches and were instructed to lift their fingers once rising indicators reached the target line (at 800ms). During Stop trials, the indicators stopped prematurely before the target and participants were instructed to inhibit the prepared finger lifts. Transcranial magnetic stimulation was delivered during response withholding (Go trials) and response inhibition (Stop trials). Arrows indicate stimulation timepoints.

The task was primarily composed of Go trials (67%, 320 trials), where participants released both switches (index finger abduction) to intercept the rising indicators with a stationary target line at 800 ms. During Stop trials (33%, 160 trials), both indicators stopped before reaching the target, cueing participants to inhibit the prepared bimanual movement. The 2:1 ratio ensured Go trials were the primed response. As is common practice^20^, an adaptive staircase algorithm was incorporated into Stop trials, dynamically changing the stop-signal delay (SSD) based on trial performance to keep task difficulty consistent across participants (∼50% success). Visual feedback was displayed at the completion of each trial to encourage accurate performance. Please see Supplementary Material 1 for more detailed information on the task.

### Recording procedure

#### Electromyography

Surface EMG (Delsys, Natick, US) was recorded over the first-dorsal interosseous (FDI) muscle of each hand as the primary agonists. The ground electrode was placed on the bony prominence of either elbow. Electrode signals were amplified, filtered (20–450 Hz), and sampled at 2 kHz (Cambridge Electronic Designs 1401, Cambridge, UK) for offline analysis with Signal (CED, v. 6.05a) and custom MATLAB software (R2019b, The MathWorks). EMG was recorded for 1 s (the length of a trial) from when the indicators started to rise.

#### Transcranial Magnetic Stimulation

Single-pulse TMS was applied to the M1 contralateral to the non-dominant (HC) or affected (PwPD) hand using a figure-of-eight D70^2^ coil and Magstim200^2^ unit (Magstim, Dyfed, UK). These hands were chosen to maximise the likelihood of detecting relevant effects, with the affected side in PwPD more susceptible to disease-related changes, and the non-dominant hand in HCs a more valid comparison. The coil was positioned tangentially to the head with the handle directed posteriorly at a 45° angle to the midline of the head, inducing a current directed posterior to anterior in the underlying cortical tissue^38^. The optimal coil position was identified for the contralateral FDI using a suprathreshold intensity. Coil placement was recorded and monitored using Brainsight® neuro-navigation software (Rogue Research, Cambridge, US). Each participant performed a practice block of 10-50 Go trials where TMS was applied 200 ms before the target to determine an intensity for the experimental blocks which consistently produced 200-400 µV MEPs.^31^

On Go trials, TMS was delivered 22 times at each of 50 ms intervals ranging from −350 to −100 ms relative to the target (132 total) to capture the CME profile during response withholding (Fig. 1). On Stop trials, stimulation was delivered at 150, 190 and 230 ms (40 times each) following the stop-signal.

### Dependent measures

#### Task performance

Lift-times on unstimulated Go trials were recorded for each index finger and the mean calculated after trimming outliers (± 3 SDs).^31^ The following trials were excluded from all analyses: stimulated trials where the coil deviated more than 3 mm/5° from the target position, and root-mean-squared (rms)EMG exceeding 15 µV 300–400 ms into the trial. Coefficient of variation was calculated as a measure of relative lift-time variability for each side. Stop signal reaction time (SSRT) was calculated using the integration method,^39^ as an index of inhibitory control.

#### Muscle bursting

EMG data processing was performed using MATLAB (MathWorks, R2022a). As per the study aims, ineffective bursts of muscle activity (i.e. those that did not trigger a lift response) were identified on a trial-by-trial basis across Go and successful Stop trials. Muscle bursts were automatically detected using a single-threshold algorithm^4^ when smoothed rectified EMG surpassed 15 SD of the baseline (0-400 ms rmsEMG) and remained elevated for ≥5 ms. Bursts separated by <15 ms were merged. Detected bursts were manually reviewed on a trial-by-trial basis. Stimulus artifacts, MEPs, erroneous or unclear bursts (e.g., no clear separation with the main burst) and contaminated bursts (interference with the MEP) were excluded from subsequent analyses. For each burst, onset/offset, duration and peak amplitude were recorded.

On Go trials, the main burst generating the lift response was identified as the last burst with an onset before the recorded switch release. Accordingly, *premature bursts* were identified as suprathreshold EMG activity prior to the main lift response. On successful Stop trials, *partial bursts* represented muscle activity which decreased in amplitude before generating sufficient force to trigger the lift response. Bursts occurring within an individualised ‘box-of-interest’ (±3 SD of trimmed Go lift-times) were classed as partial bursts, indicating preparation to respond. CancelTime was calculated as the difference in milliseconds between the SSD and peak amplitude of the partial burst (e.g., when muscle activation begins to decrease).^40^

To index burst incidence, burst trial incidence (% of traces with ≥1 burst(s)), burst frequency (mean bursts per bursting trace) and cumulative burst rate (incidence × frequency) were calculated for premature and partial bursts. All bursting measures were also recorded separately for each hand for comparison between more and less affected sides.

#### Corticomotor excitability

CME was measured via peak-to-peak MEP amplitude in the FDI of the non-dominant (HC) or affected (PD) hand. At each stimulation time, MEPs were trimmed by excluding the upper and lower 10% (if ≥10 MEPs; met in all but 7/162 HC and 18/171 PwPD data points) for a more accurate measure of centrality,^41^ as previously described.^31^ Pre-trigger muscle activity was calculated via rmsEMG 5-55 ms prior to the TMS pulse.

#### Self-report impulsivity

The Barratt Impulsiveness Scale (BIS-11)^42^ assessed trait impulsivity. Two job-related questions were excluded as participants were often retired. Therefore, total scores ranged from 28 to 112, where higher scores indicate greater impulsivity.

The Questionnaire for Impulsive-Compulsive Disorders in Parkinson’s Disease Rating Scale (QUIP-RS) recorded ICD behaviours.^43^ HCs did not respond to four questions specific to PD medication (maximum score of 96 versus 112). Total scores were converted to percentages to enable between-group comparison. Supplementary Material 1 contains more detailed information for readers unfamiliar with these two questionnaires.

### Statistical analysis

Participant demographic, cognitive and self-report impulsivity measures were compared between groups (Table 1). Kolmogorov–Smirnov tests identified any violations of normality. Non-normal data were log-transformed prior to being entered into an analysis of variance (ANOVA).

**Table 1.**
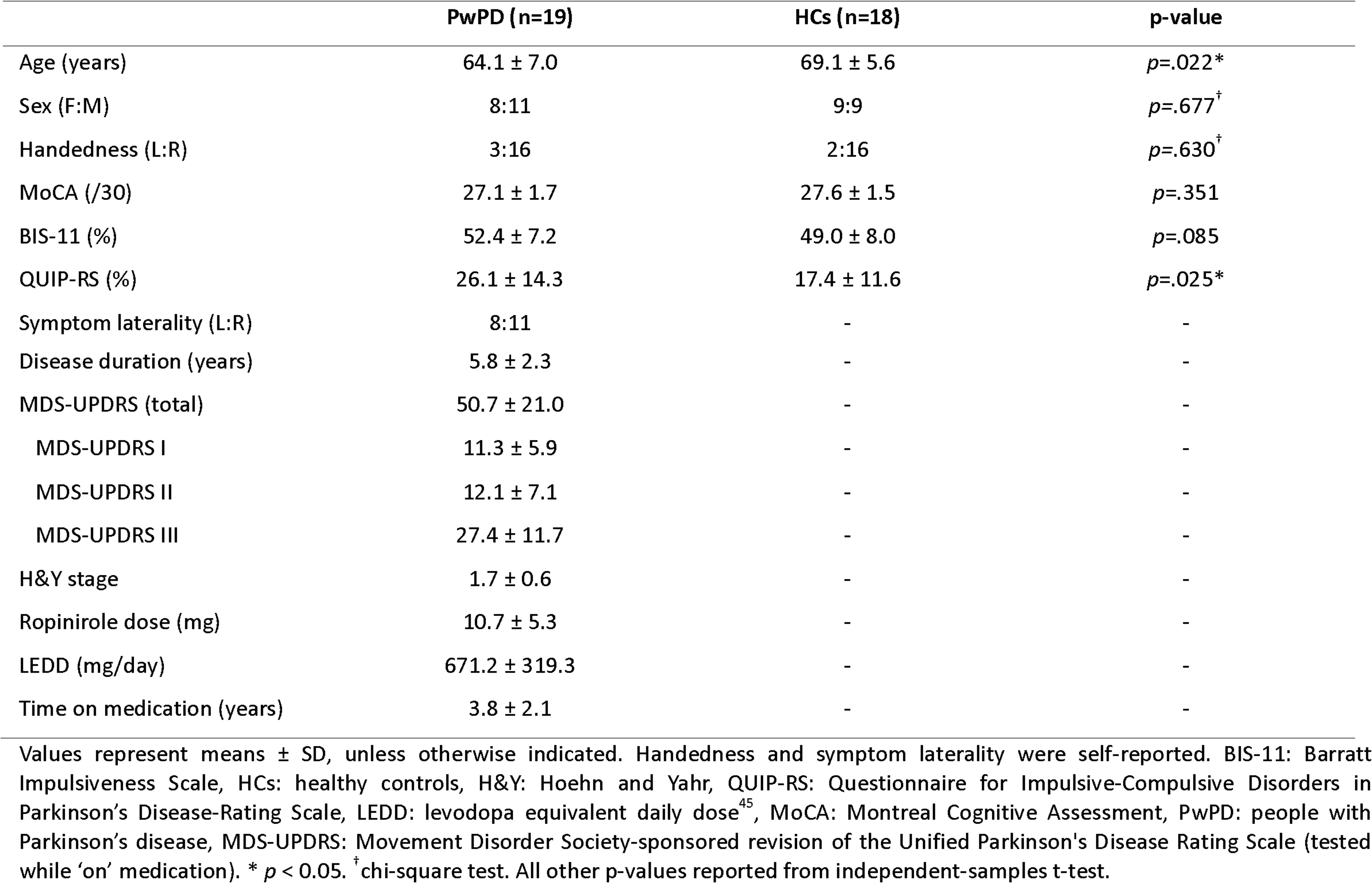
Participant demographic, cognitive and clinical variables separated by study group.

|  | PwPD (n=19) | HCS (n=18) | p-value |
| --- | --- | --- | --- |
| Age (years) | 64.1 ± 7.0 | 69.1 ± 5.6 | $p=.022^*$ |
| Sex (F:M) | 8:11 | 9:9 | $p=.677^\dagger$ |
| Handedness (L:R) | 3:16 | 2:16 | $p=.630^\dagger$ |
| MoCA (/30) | 27.1 ± 1.7 | 27.6 ± 1.5 | $p=.351$ |
| BIS-11 (%) | 52.4 ± 7.2 | 49.0 ± 8.0 | $p=.085$ |
| QUIP-RS (%) | 26.1 ± 14.3 | 17.4 ± 11.6 | $p=.025^*$ |
| Symptom laterality (L:R) | 8:11 | - | - |
| Disease duration (years) | 5.8 ± 2.3 | - | - |
| MDS-UPDRS (total) | 50.7 ± 21.0 | - | - |
| MDS-UPDRS I | 11.3 ± 5.9 | - | - |
| MDS-UPDRS II | 12.1 ± 7.1 | - | - |
| MDS-UPDRS III | 27.4 ± 11.7 | - | - |
| H&Y stage | 1.7 ± 0.6 | - | - |
| Ropinirole dose (mg) | 10.7 ± 5.3 | - | - |
| LEDD (mg/day) | 671.2 ± 319.3 | - | - |
| Time on medication (years) | 3.8 ± 2.1 | - | - |

Values represent means ± SD, unless otherwise indicated. Handedness and symptom laterality were self-reported. BIS-11: Barratt Impulsiveness Scale, HCs: healthy controls, H&Y: Hoehn and Yahr, QUIP-RS: Questionnaire for Impulsive-Compulsive Disorders in Parkinson's Disease-Rating Scale, LEDD: levodopa equivalent daily dose<sup>45</sup>, MoCA: Montreal Cognitive Assessment, PwPD: people with Parkinson's disease, MDS-UPDRS: Movement Disorder Society-sponsored revision of the Unified Parkinson's Disease Rating Scale (tested while 'on' medication). \* $p < 0.05$ . $^\dagger$ chi-square test. All other p-values reported from independent-samples t-test.

To determine whether PwPD demonstrated impaired response withholding (hypothesis 1a), lift-times (mean, coefficient of variation) were examined in a 2×2 mixed-effects ANOVA with factors Hand (more affected/non-dominant, less affected/dominant) and Group (PwPD, HC). To examine response inhibition performance (hypothesis 1b), between-group comparisons of SSRT and CancelTime were conducted using one-tailed independent-samples t-tests. CancelTime was included post-registration as a novel index of response inhibition recently validated with the ARIT.^24^ To assess the level of agreement between these indices (standard SSRT vs. novel CancelTime), a Pearson correlation analysis was conducted.

To examine muscle bursting patterns during response withholding (hypothesis 2a), the following premature burst characteristics were examined between groups using a multivariate ANOVA: burst trial incidence, burst frequency, cumulative burst rate, amplitude and duration. The same analysis was performed on partial bursts to explore muscle bursting dynamics during response inhibition (hypothesis 2b). Bursting characteristics that differed between groups were further compared between the more and less affected sides of PwPD using two-tailed paired-samples t-tests.

To examine fluctuations in CME while a response was withheld (hypothesis 3a), a 2-Group×6-Stimulation Time (−350, −300, −250, −200, −150, −100 ms relative to target) mixed ANOVA was conducted on mean MEP amplitude in Go trials. To investigate CME fluctuations while a response was inhibited (hypothesis 3b), a 2-Group×3-Stimulation Time (150, 190, 230 ms relative to stop-signal) mixed ANOVA analysed MEP amplitude on successful Stop trials. To ensure pre-trigger muscle activity did not influence CME changes, the analyses were repeated with pre-trigger rmsEMG values.

To explore potential associations between neurophysiological measures, task performance and self-reported impulsivity in PwPD (hypothesis 4), stepwise linear regression models were applied. Predictor variables were selected based on significant group comparisons. Regression models were performed on log-transformed data to meet normality and homoscedasticity assumptions.

Post-hoc comparisons were performed where necessary, with statistical significance set at α=0.05. Greenhouse-Geisser p-values are reported for violations of sphericity. Results are presented as group means ± SD.

## Results

### Demographic and clinical data

Table 1 provides a summary of demographic and clinical data, with group comparisons where applicable. PwPD were younger than HCs; therefore, age was included as a covariate in ANOVAs. Symptom severity ranged from mild to moderate (MDS-UPDRS III: 9-55)^44^ and disease duration from 1 to 9.8 years. On average, participants took ropinirole 4.3 ± 3.7 hours before the study session began. Two participants were on ropinirole as primary therapy, 16 were also on levodopa, seven on rasagiline, two on safinamide, two on entacapone, one on opicapone and one on rotigotine.

### Behavioural data

#### Lift-times

The mean lift-times of both PwPD (830.9 ± 24.2 ms) and HCs (821.5 ± 14.2 ms) were slightly after the target on Go trials (Fig. 2A). There was no main effect of Hand (*F*_1,36_=1.8, *p*=.188), or Group (*F*_1,35_=3.4, *p*=.074) or Hand × Group interaction (*F*_1,36_=3.1, *p*=.089). However, there was a main effect of Group (*F*_1,35_=7.9, *p*=.008, η_p_^2^=.185) on lift-time coefficient of variation, with PwPD exhibiting more variable lift-times than HCs (5.8 ± 1.3 vs. 4.8 ± 0.81, respectively). This effect remained when age was included as a covariate. There was no main effect of Hand (*p*=.398) or Hand × Group interaction (*p*=.609) on lift-time variability. While mean lift-times were similar between groups, increased response variability in PwPD indicates poorer control over response withholding, as hypothesised.

**Figure 2.**
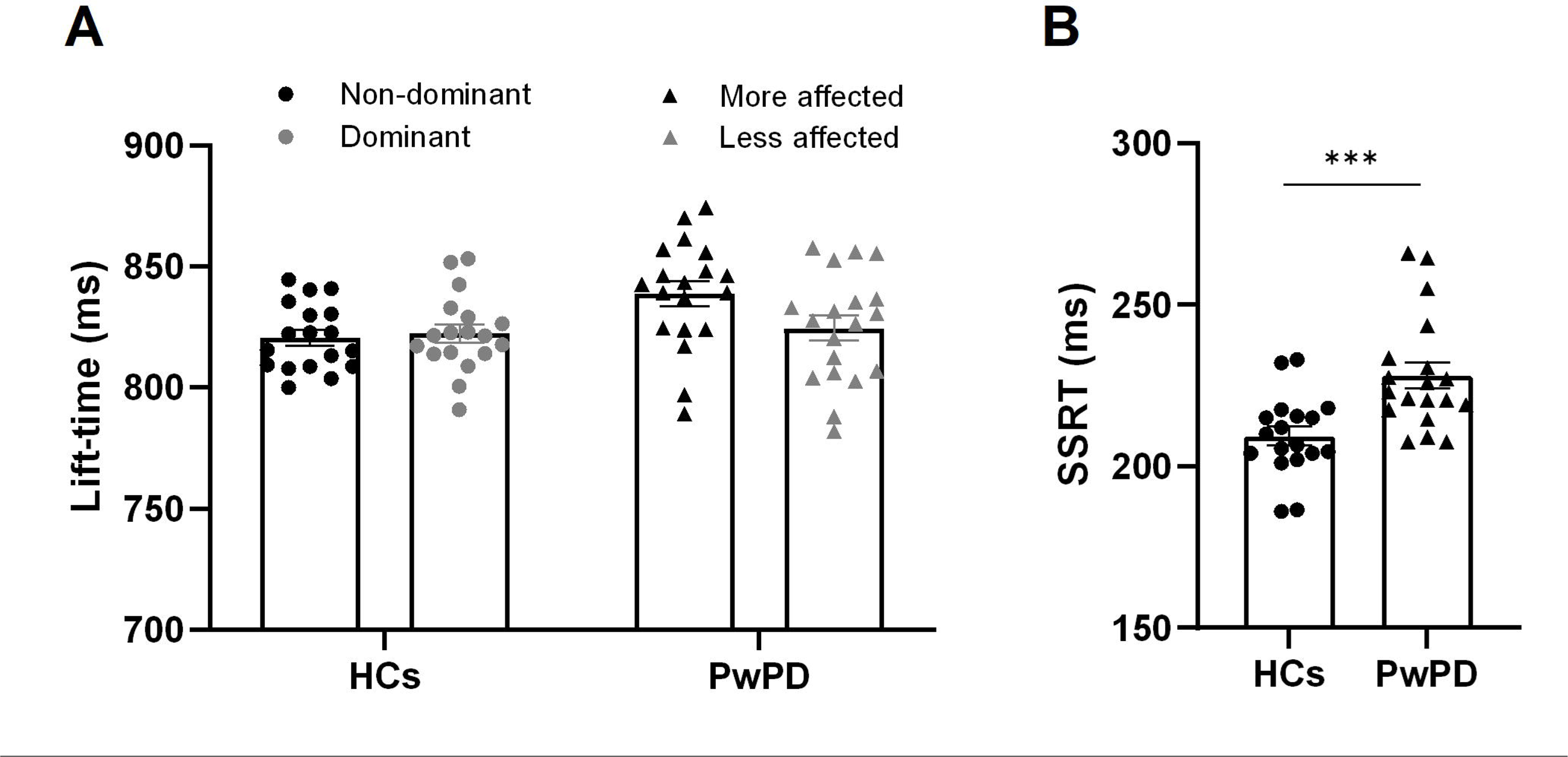
Behavioural results. Across Go trials, mean lift-times were comparable between PwPD (830.9 ± 24.2 ms) and HCs (821.5 ± 14.6 ms) (A), although lift time variability was greater in PwPD (*p*=.008). Across successful Stop trials, stop-signal reaction times (SSRT) were longer in PwPD (228.1 ± 17.6 ms) versus HCs (209.3 ± 12.4 ms, *** p < .001) (B). Values are mean ± standard error.

#### Stop Signal Reaction Time

On Stop trials, stopping success rates between 49-53% (51.2 ± 0.89%) indicate that the SSD staircasing procedure was effective. As hypothesised, PwPD demonstrated significantly longer SSRTs (228.1 ± 17.6 ms) versus HCs (209.3 ± 12.4 ms, *p*<.001, Fig. 2B), reflecting worse response inhibition. This effect was maintained after controlling for age.

### Muscle bursting

Both groups demonstrated premature muscle bursts prior to the lift response on Go trials (Fig. 3A) and partial bursts when the lift would have occurred on successful Stop trials (Fig. 4A). Mean premature burst onset ranged from 90–178 ms before the target in the HCs and 85–258 ms before the target in PwPD. Pillai’s Trace revealed a significant multivariate effect of Group on the combined bursting characteristics (incidence, frequency, rate, amplitude, duration) for both premature (*V*=.53, *F*_5,31=_6.88, *p*<.001) and partial bursts (*V*=.35, *F*_5,31=_3.38, *p*=.015). Follow-up univariate ANOVAs are described below.

**Figure 3.**
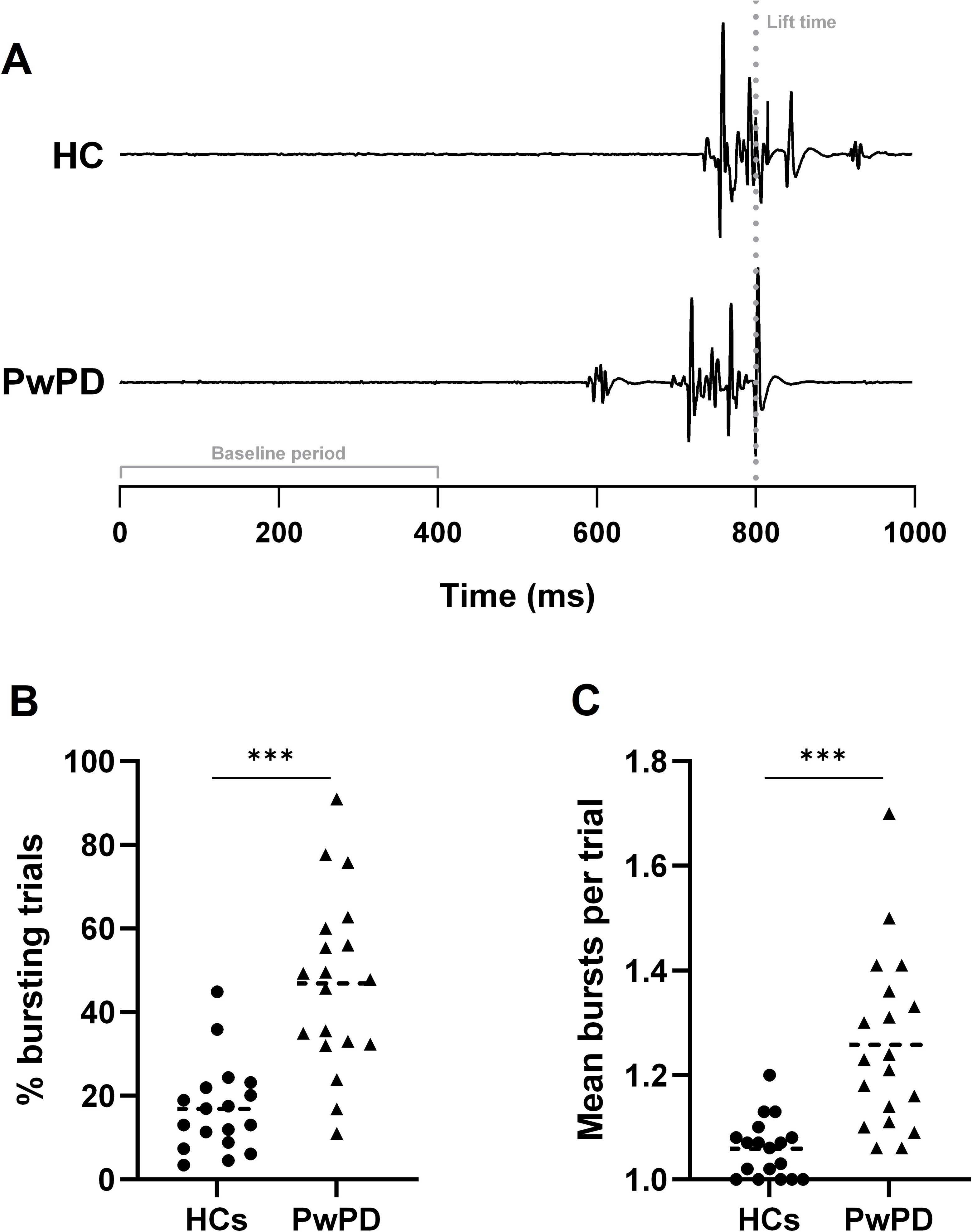
Premature bursting results. Panel A shows representative EMG traces for each group, which demonstrate a premature muscle burst (∼600 ms) prior to the main lift response in the PwPD (dotted line indicates lift-time). Panel B shows a higher percentage of Go trials with ≥1 premature burst(s) in PwPD (46.9 ± 21.0%) compared to HCs (16.9 ± 10.8%; *** *p*<.001). Panel C shows a greater frequency of bursts within these trials in PwPD (1.26 ± 0.17 vs. 1.06 ± 0.06; *** *p*<.001).

**Figure 4.**
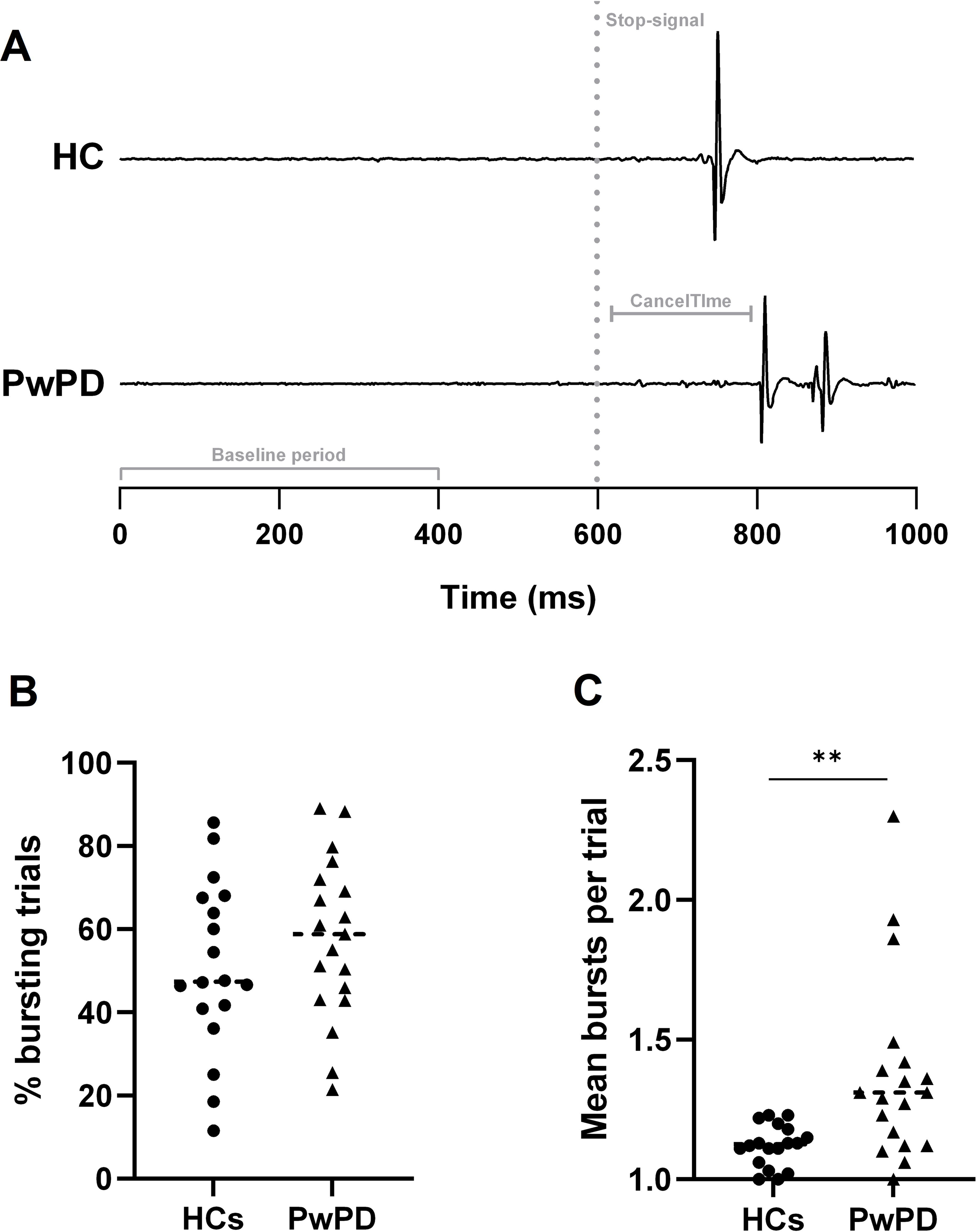
Partial bursting results. Panel A shows representative EMG traces for each group, which demonstrate partial muscles bursts in both a HC participant and PwPD following the stop-signal (dotted line) around when the response would have occurred (∼750-800 ms). Panel B shows both groups exhibited ≥1 partial muscle burst(s) in >50% of trials. Panel C shows a greater frequency of bursts within these trials in PwPD (1.37 ± 0.33 vs. 1.12 ± 0.08; *** *p*=.001).

#### Premature bursts

As hypothesised, PwPD demonstrated greater premature muscle burst activity than HCs on Go trials. There was a significant effect of Group on premature burst rate (*F*_1,35_=31.70, *p*<.001, η_p_^2^=.48), with a higher rate in PwPD (0.62 ± 0.36) compared to HCs (0.18 ± 0.13). This measure reflected a higher incidence Go trials with ≥1 premature burst(s) in PwPD (46.9 ± 21.0%) compared to HCs (16.9 ± 10.8%; *F*_1,35_=30.61, *p*<.001, η_p_^2^=.47; Fig. 3B) and a greater frequency of bursts within these trials (PwPD: 1.26 ± 0.17; HC: 1.06 ± 0.06; *F*_1,35_=26.06, *p*<.001, η_p_ ^2^=.43; Fig. 3C). There were no group differences in amplitude (*F*_1,35_=.86, *p*=.361) or duration (*F*_1,35_=.004, *p*=.953). In PwPD, premature burst incidence (*p*=.094) and frequency (*p*=.091) were comparable between the more and less affected hands.

#### Partial bursts

Both groups had a similar proportion of successful Stop trials (PwPD: 57.6 ± 19.3 %; HC: 50.9 ± 20.6 %; *p*=.279) containing ≥1 partial muscle burst(s) (Fig. 4B). However, these bursts occurred more frequently within-trials in PwPD (1.37 ± 0.33) compared to HCs (1.12 ± 0.08; *F*_1,35_=11.92, *p*=.001, η_p_^2^=.25; Fig. 4C). The main effect of Group on burst rate did not reach statistical significance (PwPD: 0.81 ± 0.37; HC: 0.58 ± 0.26; *F*_1,35_=3.83, *p*=.058). Again, there were no groups differences in amplitude (*F*_1,35_=.72, *p*=.401) or duration (*F*_1,35_=3.61, *p*=.066) and partial burst frequency was comparable between the two sides in PwPD (*p*=.359).

In accordance with our SSRT findings, CancelTime was longer in PwPD (184.5 ± 44.6 ms after the stop-signal) than HCs (160.1 ± 14.0 ms; *p=*.015), reflecting delayed inhibition latency. As expected, there was a positive correlation between CancelTime and SSRT (*r=*.617, *p*<.001).

### Corticomotor excitability

#### Response withholding

Stimulation intensity was set at 48.7 ± 8.3% to obtain baseline MEPs in the 200-400 µV range. For Go trials (Fig. 5A), CME increased in both groups as the target approached (*F*_5,175_=182.4, *p*<.001, η^2^_p_=.839). However, a Group × Stimulation Time interaction (*F*_5,175_=3.8, *p*=.003, η^2^_p_=.099) revealed that CME increased earlier in PwPD at −300 ms (207.7 ± 107.5 μV vs. 128.3 ± 82.6 μV, *p*=.021), and remained above HCs until −200 ms (494.4 ± 359.2 μV vs. 213.0 ± 140.8 μV, *p*=.003). The observed effects became more pronounced after controlling for age. PwPD additionally demonstrated higher CME than HCs at −150 ms when age was added as a covariate (885.0 ± 594.1 μV vs. 688.2 ± 654.5 μV, *p*=.027). Both groups had comparable CME values at the final timepoint (−100 ms).

**Figure 5.**
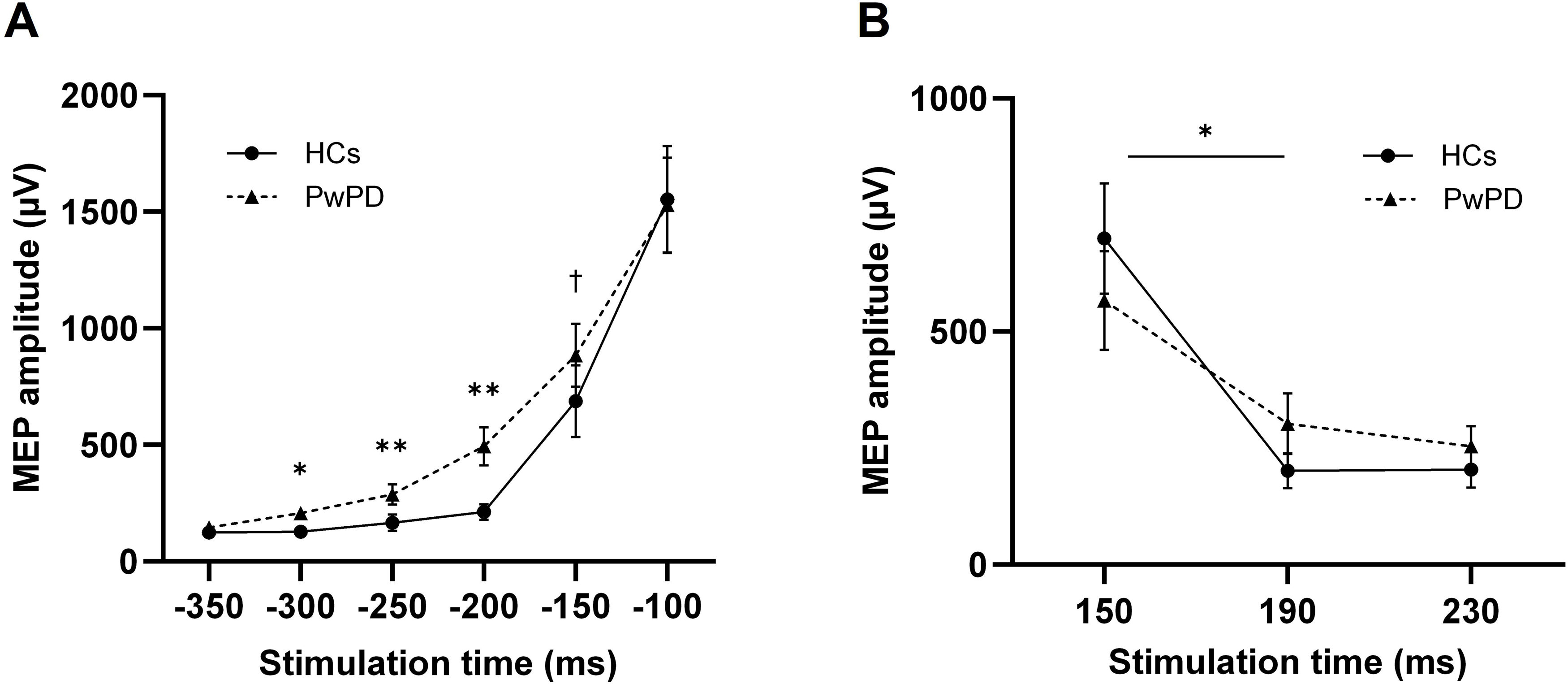
Corticomotor excitability (CME) results. In Go trials (A), while CME increased across both groups in anticipation of the target, PwPD demonstrated an earlier increase in CME from −300 ms to −150 ms. Between-groups differences at each timepoint are indicated with * *p* < 0.05; ** *p* < 0.01; † *p* < 0.05 with age as a covariate. In successful Stop trials (B), while both groups demonstrated a reduction in CME, PwPD exhibited a smaller drop in CME from 150 ms to 190 ms (* *p* = .025). Values are mean ± standard error.

There was no main effect of Group (*F*_1,35_=.013, *p*=.908) or Group × Stimulation Time interaction (*F*_5,175_=1.84, *p*=.139) for pre-trigger rmsEMG on Go trials. There was a main effect of Stimulation Time (*F*_5,175_ = 59.07, *p*<.001, η^2^_p_ =.628), with rmsEMG rising 200 ms prior to the target in both groups.

#### Response inhibition

For MEP amplitude on successful Stop trials, there was a main effect of Stimulation Time (*F*_2,70_=44.9, *p*<.001, η^2^_p_ =.562), with an overall reduction in MEP amplitude between 150 ms (631.3 ± 479.9 μV) and both 190 ms (252.7 ± 233.6 μV, *p*<.001) and 230 ms (229.3 ± 177.1 μV, *p*<.001) following the stop-signal. There was no main effect of Group (*F*_1,35_=.002, *p*=.961), however, there was a Group × Stimulation Time interaction (*F*_2,70_=4.3, *p*=.025, η^2^_p_=.109), with PwPD demonstrating a smaller drop in MEP amplitude between 150–190 ms (264.8 ± 338.6 μV) following the stop-signal compared to HCs (498.7 ± 413.0 μV, *p*=.034; Fig. 5B). The above effects mostly remained consistent when age was included as a covariate, although the Group × Stimulation Time interaction trended towards significance (*p*=.051).

For pre-trigger rmsEMG, there was a main effect of Stimulation Time (*F*_2,70_=73.91, *p*<.001, η^2^_p_ =.679), with an overall reduction at 230 ms (17.2 ± 18.1 μV) compared to 150 ms (52.5 ± 59.8 μV, *p<*.001) and 190 ms (57.4 ± 65.1 μV, *p<*.001). There was also a Group × Stimulation Time interaction (*F*_2,70_=4.04, *p*=.022), with higher rmsEMG values in HCs than PwPD at 190 ms (57.4 ± 51.3 μV vs. 47.9 ± 67.9 μV, *p*=.02) and 230 ms (74.3 ± 66.9 μV vs. 41.4 ± 60.8 μV, *p*=.009). Crucially, this interaction revealed an inverse relationship between rmsEMG and CME changes, with HCs exhibiting *higher* rmsEMG alongside a larger *decrease* in MEP amplitude compared to PwPD. Therefore, there was no group difference in pre-trigger muscle activity across 150–190 ms that could account for the difference in MEP amplitudes.

### Regression models

#### Predicting response withholding

A stepwise linear regression was conducted to model response withholding performance, using lift-time variability in Go trials as the outcome variable. Predictor variables were premature burst rate, mean CME across the timepoints showing increased excitability in PwPD (−300 ms, −250 ms, and −200 ms), and their interaction.

Premature burst rate interacted with CME to predict lift-time variability (F_1,17_=5.59, *p*=.03), accounting for 20% of the variance (R²=.248, adjusted R²=.203). A simple slopes analysis indicated that increased premature burst rates combined with high CME levels were associated with greater lift-time variability (*p*=.038, *r*=0.836).

#### Predicting response inhibition

Two stepwise linear regressions were conducted to model response inhibition performance, using SSRT and CancelTime as outcome variables. Predictor variables were partial burst frequency, CME difference score (150–190 ms), and their interaction.

In the SSRT model, none of the predictor variables met the statistical entry criteria (*p*<.05) for inclusion. However, partial burst frequency emerged as a significant predictor of CancelTime (F_1,17_=38.65, *p*<.001. β=.833), accounting for 67.7% of the variance (R²=.695, adjusted R²=.677). Specifically, a 10% increase in partial burst frequency was associated with a ∼8.3% increase in CancelTime.

#### Predicting self-report impulsivity

Two stepwise linear regressions were conducted to model self-reported impulsivity in PwPD, measured via the BIS-11 and QUIP-RS. As per the previous models for task performance, neurophysiological predictor variables included premature burst rate and mean CME in Go trials, and partial burst frequency, CME difference score and CancelTime in Stop trials, as well as multiple interaction terms capturing within- and between-trial dynamics:

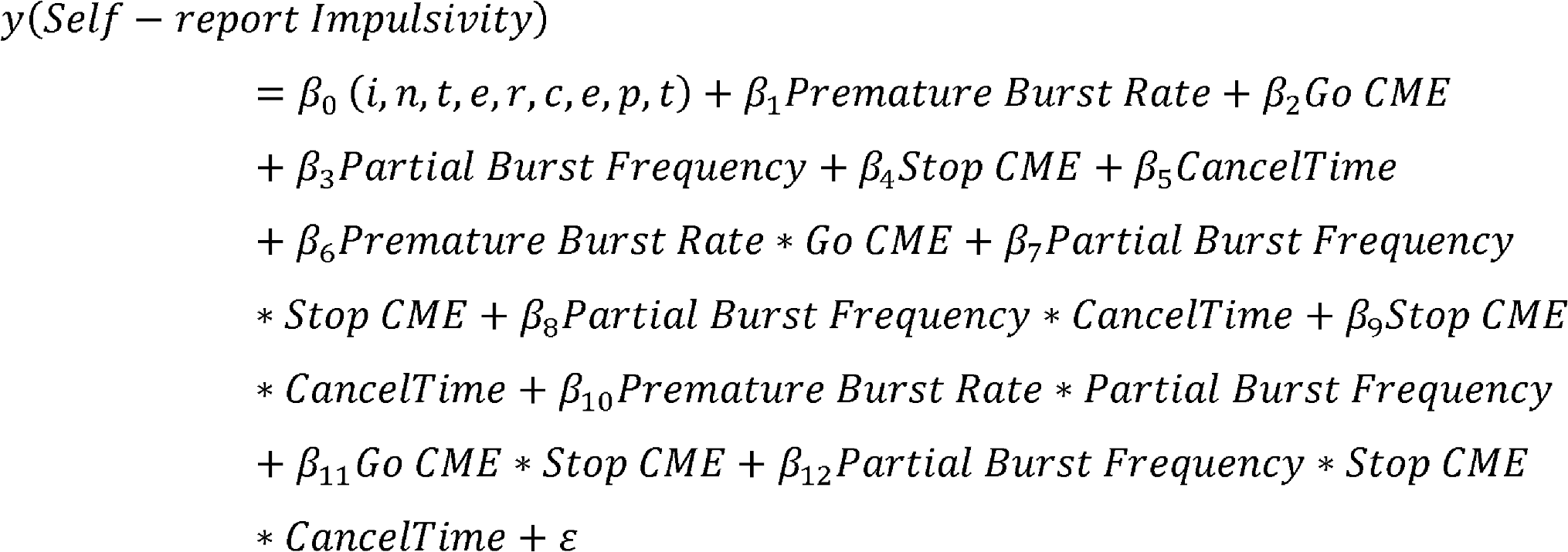

None of the predictor variables met the statistical entry criteria (*p*<.05) for inclusion in the final BIS-11 or QUIP-RS models. To check the validity of the reported QUIP-RS scores, we examined whether a previously reported association between response inhibition performance and clinical impulsivity could be replicated^22^ by incorporating SSRT into the existing model. Indeed, the resulting model indicated that SSRT was a significant predictor of QUIP-RS score (*F*_1,17_=9.22, *p*=.007, β=4.859), accounting for 31.3% of the variance (*R*²=.352, adjusted *R*²=.313). Specifically, a 10% increase in SSRT was associated with a ∼48.6% increase in QUIP-RS score.

#### Exploratory post-hoc analyses

Follow-up stepwise regression analyses tested for potential relationships between clinical variables (disease duration, MDS-UPDRS total score, Hoehn and Yahr stage, ropinirole dose, LEDD and time on medication) and behavioural/neurophysiological measures that showed group differences (premature burst rate, Go CME, lift-time variability, partial burst frequency, Stop CME, CancelTime and SSRT). These analyses revealed a significant interaction between symptom severity (MDS-UPDRS total) and overall medication dose (LEDD) in predicting several measures related to response inhibition, including partial burst frequency (*F*_1,17_=7.25, *p*=.015, adjusted *R*²=.258), CancelTime (*F*_1,17_=4.70, *p*=.045, adjusted *R*²=.171) and SSRT (*F*_1,17_=4.89, *p*=.041, adjusted *R*²=.178). A simple slopes analysis indicated that for those with greater symptom severity, higher medication doses were associated with increased partial bursting (*p*=.016, *r*=.895) and a trend toward longer CancelTime values (*p*=.086, *r*=.750).

## Discussion

The present study generated several novel findings regarding the mechanisms underlying impaired impulse control in PwPD. We identified objective neurophysiological measures sensitive to deficits in response withholding and response inhibition in PD, which were associated with a more severe clinical profile. Compared to healthy older adults, PwPD exhibited both a higher rate of dysfunctional muscle bursts and an early transient increase in CME during Go trials, manifesting as more variable lift responses. PwPD also demonstrated a smaller reduction in CME and a higher frequency of muscle bursts on Stop trials which led to impaired response inhibition performance, that was itself predictive of more clinically problematic impulsive behaviours. We provide evidence that proactive and reactive inhibitory processes are compromised in PD at both a behavioural and neurophysiological level. Importantly, difficulties with general motor control in PD and background muscle activity were unable to account for our novel neurophysiological results. We discuss the theoretical and clinical implications of our findings below, including whether the changes observed in our PD sample may reflect the early effects of DA medication on the inhibitory control network, triggering increased impulsive action before the emergence of overt ICD symptoms.

### Theoretical implications

#### Response withholding

Consistent with previous research,^15,46,47^ mean lift-times were comparable between PwPD and healthy older adults. Instead, we confirmed an increased variability in ARIT lift responses for PwPD,^47^ which has previously been shown on externally cued tasks.^48–51^ Our results showed that a higher rate of premature muscle bursts (both within and across trials) at high CME levels in PwPD was associated with the increased variability in lift responses. Interestingly, however, the amplitude and duration of these premature bursts did not differ between groups. Therefore, we propose their occurrence alone serves as a marker of the state of the inhibitory control system, signalling less reliable response withholding resulting from impaired proactive inhibitory control in PwPD.^52^ The predictive capability of muscle bursts highlights the sensitivity of the measure to fluctuations in task performance, offering a more nuanced understanding of inhibitory control function.

While healthy older adults maintained a stable inhibitory state (as per^30–32^) until ∼150 ms before the target—at which point MEP amplitudes increased above baseline—PwPD demonstrated an early and transient rise in excitability above HCs ∼300 ms before the target. This shift in CME preceded the onset of premature muscle bursts (Fig. 6), perhaps reflecting impairment in preparatory suppression,^18,19,53^ whereby elevated CME propagates through the motor system, triggering muscle bursting and thereby increasing response variability. Such impairment could be mediated via dysfunctional activity in GABAergic inhibitory networks within M1,^54–56^ a potential area for future research.

**Figure 6.**
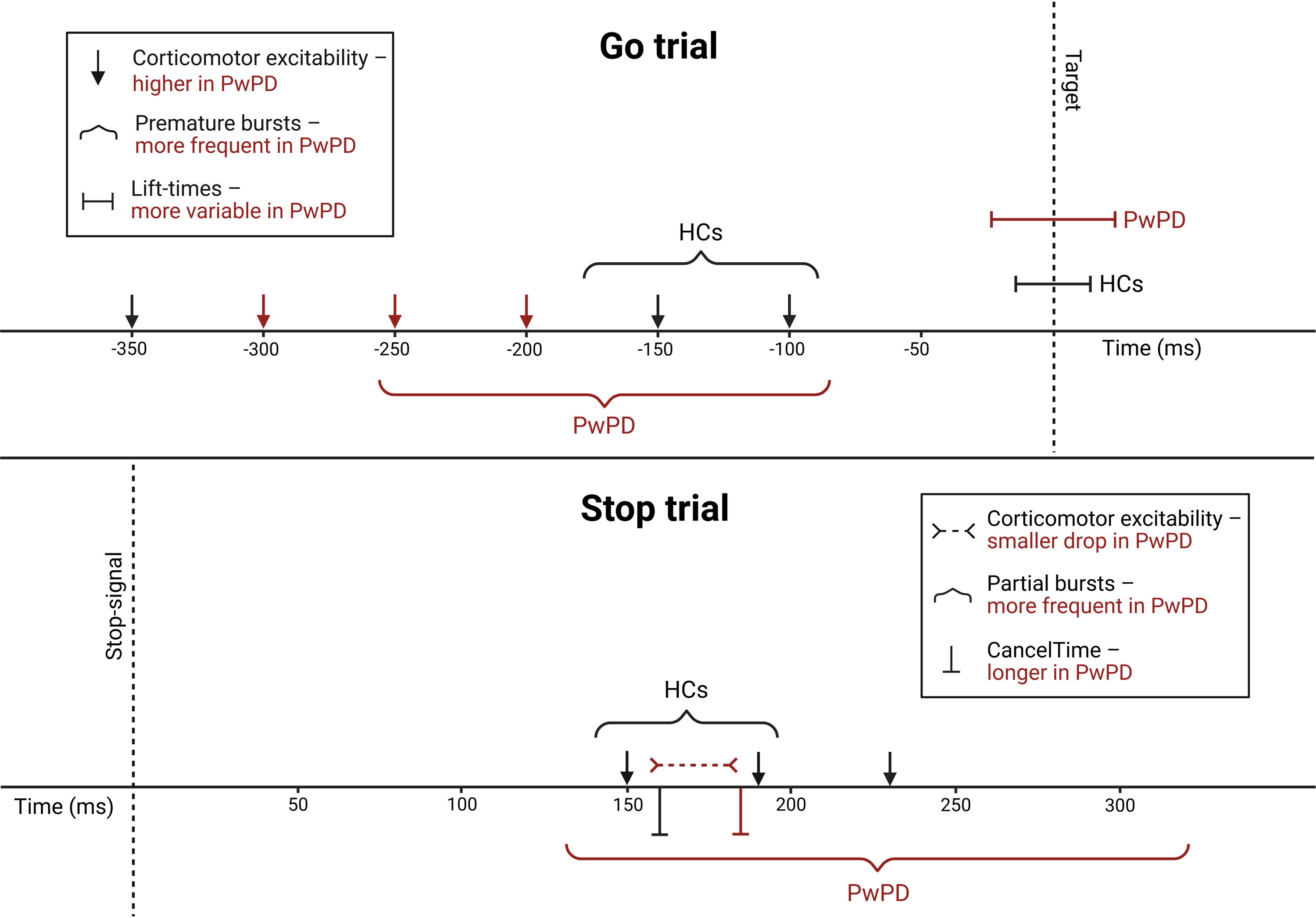
Results summary. A schematic of the relative timings of the corticomotor excitability, muscle bursting and behavioural results across Go and successful Stop trials. Group differences between PwPD and HCs are indicated in red.

#### Response inhibition

PwPD demonstrated impaired inhibition of the anticipated response at both behavioural and neurophysiological levels. In accordance with previous research,^11,12,15,46^ our PwPD group showed longer SSRTs compared to HCs, however, we are the first to show that the related measure of CancelTime was also longer in PwPD. A greater number of partial bursts within individual trials was associated with longer CancelTime values, indicative of fragmented motor suppression. Partial burst frequency is therefore a potential trial-by-trial indicator of reactive response inhibition, reflecting how ineffective engagement of the inhibitory control network in PD allows multiple bursts of muscle activity to emerge, thus delaying complete inhibition.

While a greater number of partial bursts occurred during response inhibition trials, the overall number of successful Stop trials exhibiting partial bursts was comparable between PwPD and HCs. This is not entirely surprising, as partial bursts have been observed in healthy adults across the stop-signal task^24,40^ and ARIT,^24,25^ although to a lesser extent. The increased prevalence (∼50%) in our study may reflect age-related slowing of inhibition (see^57^ for a meta-analysis), potentially leading to greater difficulty in suppressing motor activity. Therefore, partial burst prevalence may not serve as a sensitive marker of PD-specific inhibitory deficits but rather reflect broader age-related declines in motor control.

While healthy older adults dynamically suppressed CME during successful stopping, consistent with findings in younger cohorts,^31,33^ PwPD demonstrated a blunted inhibitory response that failed to sufficiently clamp down on CME in a timely manner. As pre-trigger muscle activity did not mirror the observed MEP changes, the attenuated CME suppression in PD reflects processes upstream of the alpha-motoneuron pool, likely within dopaminergic cortico-basal ganglia networks which show well-documented impairments in PD.^58–60^ For instance, successful response inhibition is thought to originate via activation of the right inferior frontal gyrus and pre-supplementary motor area^61–63^ which are both PFC regions impacted by PD.^64,65^ It is plausible that the results in the current study are mediated by deficits in top-down inhibitory signalling^66^ from disruption in these frontal regions, leading to disinhibition of the motor system in PwPD, demonstrated by reduced CME suppression, increased partial bursting and impaired stopping performance (Fig. 6). Future neuroimaging research could more clearly elucidate how motor-level and prefrontal cognitive-level contributions interact to shape inhibitory control in PwPD.

Interestingly, while our partial bursting and CME measures appear sensitive to impaired task-related response inhibition, neither emerged as significant predictors of self-reported impulsivity. While these neurophysiological markers appear to effectively capture motor impulsivity, this may not (yet) directly translate to real-world impulsive behaviours. The predictive link between clinical severity and partial bursting suggests potential for translation to everyday behaviour in the future. Nevertheless, currently only SSRT emerged as a predictor of clinical impulsivity (via QUIP-RS), with longer SSRTs linked with increased ICD severity in PwPD taking DAs, which aligns with previous research.^22^ Of note, while neither of our neurophysiological markers were linked with SSRT, more frequent partial bursts were associated with longer CancelTime values. It appears, therefore, that CancelTime and SSRT reflect distinct aspects of inhibitory control, despite being strongly correlated. Our findings highlight a potential dissociation between behavioural and neurophysiological measures of response inhibition, with CancelTime providing a granular, motor-level index of inhibitory efficiency, while SSRT captures broader impulse control behaviour (see Supplementary Material 2 for in-depth discussion). This underscores the importance of integrating both behavioural and neurophysiological measures when assessing impulse control, as they can capture distinct yet complementary aspects of the inhibition process.

### Clinical implications

The behavioural and neurophysiological changes observed in our PD sample may reflect the relatively early effects of DA medication on the inhibitory control network, with the average time on ropinirole only 3.8 ± 2.1 years. Unfortunately, the specific effects of dopamine medication on impulse control behaviour in PwPD are mixed and likely occur on an individualised level according to patient characteristics (e.g., disease status, baseline behaviour), medication profiles and the employed task.^9^ Given the diverse medication profiles in our PD sample, we cannot isolate the specific effects of ropinirole from those of concurrent medications. For instance, the interacting effects of levodopa with dopamine agonists have been linked with hyper-responsive dopaminergic neuron firing to rewarding stimuli, potentially fostering impulsive behaviours.^67^ Indeed, our post-hoc regressions revealed that symptom severity interacted with overall medication dose to predict neurophysiological measures of response inhibition (partial bursting and CancelTime). This extends findings from a meta-analysis,^12^ whereby longer disease duration predicted greater impulsive action deficits in PwPD on medication. Interestingly, while higher ropinirole doses have specifically been linked to impulsive behaviour,^68^ no such association was found in the current study, suggesting that overall dopamine levels had a greater impact on impulse control performance.

This link between a more severe clinical profile and neurophysiological markers of impaired inhibition suggests that some individuals were shifted beyond the optimal dopamine range, particularly in circuitry that remains relatively intact.^69^ Those with more progressed pathology may require higher doses to manage motor symptoms, at the cost of shifting mesocorticolimbic networks implicated in cognitive/limbic control into a hyperdopaminergic state, resulting in neurophysiological/behavioural impairment.^70,71^ Clinically, these findings underscore the importance of individualised treatment strategies that consider the potential non-motor side effects of dopaminergic therapy. Neurophysiological markers such as partial bursting and CancelTime may serve as early indicators of dopamine overdose effects on impulse control before the emergence of overt behavioural symptoms like ICDs.

Importantly, despite a range of symptom and medication profiles, we observed robust group effects across the applied behavioural and neurophysiological measures, reinforcing the presence of inhibitory control deficits across a heterogeneous PD sample. However, without comparison to PwPD who are medication-naïve (de novo) or temporarily withdrawn from dopaminergic treatment, we cannot disentangle medication effects from pathology-related inhibitory dysfunction. Further exploration of these measures in de novo PD would help determine whether the neurophysiological changes observed in the current study emerge prior to behavioural impairments early in the disease course,^72^ potentially serving as biomarkers for future impulse control problems.

### Potential limitations

The present study primarily involved participants in mild-to-moderate stages of PD due to task demands requiring fine motor movement. While our sample included a range of symptom severities (MDS-UPDRS III: 9–55), participants were limited to Hoehn and Yahr stages ≤2.5 and disease durations <10 years. Notably, two participants were excluded due to more severe motor symptoms invalidating task performance, highlighting the challenge of independently assessing response inhibition in later disease stages. Considering the observed link between symptom severity and markers of inhibition, future work should consider adapting response inhibition tasks to accommodate more advanced motor impairments to better understand how inhibitory control networks are affected by disease progression.

It is also worth considering the confounding effects of age-related cognitive decline. Although participants were screened for substantial cognitive impairment, early-stage cognitive decline may have influenced impulse control performance. As HCs were older than PwPD, age was included as a covariate in the applied analyses to account for potential age-related slowing of cognitive-motor inhibition. Importantly, this adjustment had minimal impact on the results, strengthening the robustness of our conclusions. Please see Supplementary Material 2 for methodologically detailed considerations specific to future studies investigating premature and partial muscle bursts in PwPD.

## Conclusion

The current study identified objective neurophysiological measures sensitive to impulse control difficulties in PD. Our results corroborate previous literature demonstrating impaired response inhibition in PwPD, alongside novel results showing reduced modulation of muscle activity and CME. Our findings further extend the literature by revealing evidence of premature neuromuscular activation during response withholding, resulting in more variable behavioural responses. Collectively, the current study provides neurophysiological evidence of widespread deficits across both proactive and reactive inhibitory processes in PwPD. These changes manifested as impaired task performance and were associated with a more severe clinical profile. Further investigation into the predictive capability of these objective measures for the development of ICD symptoms will help establish their potential as a valuable tool in clinical practice.

## Acknowledgements

The authors gratefully acknowledge the commitment of participants and caregivers who contributed their time to this study. The authors would also like to acknowledge Parkinson’s UK, Birmingham 1000 Elders, and the NIHR for their support with recruitment, and Brogan Ling and Joshua Pearson for assistance with data collection.

## Data availability

The anonymised datasets analysed in the current study are available from the corresponding author upon reasonable request. Access will be granted in accordance with institutional guidelines and subject to ethical approval where required.

## Supplementary material

Supplementary materials are available at *Brain* online.

