## Supplementary Material for "Muscle bursting and corticomotor excitability index impaired impulse control in Parkinson’s disease"

**Supplementary material 1 – Methodology details**

**Participants**

Participants performed the task on two separate days as part of the larger pre-registered study. In the first session, electroencephalography (EEG) was recorded, and on the second day TMS was applied, as described here. All participants were therefore familiar with the task, having completed it in a prior session. Of note, the larger study isolates the effects of PD versus medication by comparing HCs, de novo (medication-naïve) PwPD and PwPD taking ropinirole. Here, we have reported comparisons with HCs and PwPD taking ropinirole only.

Following data collection, two PwPD were excluded due to outlying behavioural values (LT/SSRT identified via the box-plot method). One PwPD and one HC were excluded due to low signal-to-noise ratio in the EMG. One HC was excluded due to unreliable baseline MEPs.

**Behavioural task**

Participants were seated ~0.6 m in front of a computer monitor displaying two vertically oriented indicators (18 cm in length, 2 cm in width, 2 cm apart) (Fig. 1). Switch ‘up/down’ state was precisely recorded (<1 ms) through an Arduino (Uno; Arduino.cc) and synchronised to the display through an analogue-digital USB interface (NI-DAQmx9.7; National Instruments). Participants rested their forearms on a table, positioned mid-way between supination and pronation. Go trials were deemed successful, displaying *Success – hit target* and turning the target green, if both indicators were within 50 ms of the target. This tolerance was adapted from previous work^1^ to ensure suitability for PD, who may experience delayed processing speeds^2^. If the indicators were outside the set tolerance, *Missed target* was presented, and the target line turned red. If no lift response was registered, then *No response* was displayed, and this trial was excluded from analyses. Following Stop trials, a green target and *Stop successful* were presented if participants held their fingers on the switches till the end of the trial, while a red target and *Stop failed* indicated failure to inhibit the lift response.

Visual feedback was also displayed at the end of each experimental block, displaying RT, Go accuracy and Stop accuracy plots. To prevent strategic slowing, participants were explicitly instructed not to delay Go responses, but to lift their fingers at the target line as accurately as possible and this was monitored via visual feedback throughout the task. To maintain clean EMG recordings, participants were asked to rest their fingers onto the switches at the beginning of each trial and activate their muscles only when lifting their fingers. Experimenters visually monitored the background EMG activity and reminded participants to relax their fingers if unwanted muscle activity was detected. The Arduino device triggered both the Magstim 200^2^ and Signal software to synchronise the stimulation timing.

**Dependent measures**

**SSRT integration method**

Trimmed lift-times averaged across side for Go trials were rank ordered and the nth lift-time selected, with n obtained by multiplying the number of lift-times by the probability of a response on a Stop trial^3^. The time at which the staircase procedure stopped the indicators to achieve 50% success (*staircased* SSD) was subtracted from the nth lift-time.

**Self-report questionnaires**

The BIS-11^5^ was used as an index of trait impulsivity, reflecting an individual’s tendency to act on impulses rather than engage in considered decision-making. The BIS-11 is a 30-item questionnaire designed to assess impulsivity across three broad domains: attentional impulsiveness (difficulty focusing and shifting attention), motor impulsiveness (acting without thinking), and non-planning impulsiveness (lack of future-oriented thinking). Participants rate each item on a 4-point Likert scale (1=rarely/never, 4=almost always).

The QUIP-RS is a validated 28-item self-report measure for ICD behaviours in PD, covering four primary ICDs—pathological gambling, compulsive buying, hypersexuality, and binge eating—as well as three related behaviours: punding, hobbyism, and dopamine dysregulation syndrome. Each item is rated on a 5-point Likert scale (0=never, 4=very often), with higher scores indicative of greater ICD symptomology.

**EMG considerations**

There are several considerations worth noting for future EMG studies investigating premature and partial muscle bursts in PwPD. EMG data quality was variable, particularly in PwPD, with the signal often weakening as the task progressed. This decline may have been due to the EMG electrodes drying out as the task progressed, alongside muscle weakness and/or fatigue characteristic of PD, as it was often most pronounced in the affected side. It is notable then that, although the EMG signal was weaker in PwPD, making it more challenging to detect muscle bursting activity, we still observed increased FDI muscle bursting within this group. However, muscle fatigue/weakness may have led to the recruitment of alternate muscles not recorded. Recording activity from compensatory muscles, including the extensor indicis and wrist muscles, could provide valuable insights into inhibitory control of the broader motor network in PD. Despite these potential limitations, EMG proved to be a sensitive tool for assessing inhibitory control, indicating that premature and partial muscle bursts are robust markers for future use in this clinical population.

This study used a specific range (3SD of trimmed lift-times) to accurately identify partial bursts where a response would have occurred in successful Stop trials. However, as in Go trials, premature bursts may also occur before the stop-signal while the prepared response is being withheld. The dynamic stop-signal staircase algorithm, continually adjusting the timing of response withholding versus response inhibition, prevented reliable classification of premature muscle bursts in Stop trials. Although not the focus of the current study, future research using a fixed, staircased SSD could provide a more robust measure of motor impulsivity by capturing both premature and subsequent partial bursts on Stop trials. This approach would help determine whether these two burst types frequently co-occur, linking impaired response withholding and inhibition within the same trial.

**Pre-trigger EMG**

Unlike much of the existing research using MEP amplitude to assess CME, we did not exclude trials based on EMG activity immediately before the TMS pulse, as these timepoints necessarily overlapped with the expected timing of muscle bursts. Therefore, automatically removing trials from the CME analysis which contained potential muscle bursts of primary interest would have weakened our ability to link CME fluctuations and muscle bursting (hypothesis 3). We instead rejected trials which showed high EMG activity (>15 μV) 300-400 ms into the trial, prior to any expected bursting activity, and compared pre-trigger muscle activity patterns in accordance with the CME analysis. While pre-trigger rmsEMG values rose in HCs during response inhibition—likely reflecting the stronger EMG signal in this group alongside the high incidence of trials with partial bursts—these changes did not correspond with the observed fluctuations in MEP amplitude. Future research should consider the potential limitation of applying traditional, ‘blind’ pre-trigger rmsEMG exclusion criteria when integrating CME and muscle burst data. Such thresholds may inadvertently exclude trials containing partial bursts, potentially masking interesting effects between muscle-level activity and CME.

**Supplementary material 2 – SSRT vs CancelTime comparison**

SSRT provides an estimate of stopping performance based on average behavioural data (e.g., lift-times, SSD), derived from the 'horse race model' of action inhibition.^6^ This model posits that independent go and stop processes compete to reach a threshold, with the faster process determining whether a response is executed or inhibited. However, this independence assumption is not always met in empirical data,^7,8^ with evidence that stopping may influence the speed of the go process, particularly in tasks like the ARIT.^9^ In practice, SSRT is reduced by proactive slowing, whereby participants slow down go responses in anticipation of possibly needing to stop.^10^ Despite these theoretical limitations, longer SSRTs have been consistently associated with increased impulsivity across both clinical^11-14^ and non-clinical populations.^15^ This association was further supported in the current study. Therefore, SSRT may serve as a broader measure of inhibitory control, relating more directly to real-world impulsive behaviours.

In contrast, CancelTime is a more direct measure of the physiological inhibitory process. CancelTime directly indexes stopping latency from muscle activity, capturing inhibition at the point where covert muscle activation begins to decrease on an individual-trial basis. Therefore, CancelTime may primarily reflect an active inhibition mechanism that blocks ongoing motor activity (e.g.,^16^) which may not (yet) translate to broader impulsivity traits. However, it is important to note that CancelTime was measured using the peak of the earliest partial burst identified within each trial. PwPD exhibited a greater number of partial bursts within-trials, indicative of incomplete inhibition. As a result, the CancelTime values reported in this study may underestimate the duration of the inhibitory process in PwPD. Future research aimed at identifying the most robust method for calculating CancelTime in the presence of multiple partial bursts may yield a more accurate neurophysiological measure of inhibition. Nevertheless, by isolating the neuromuscular mechanisms underlying response inhibition from potential experimental variations (e.g., pressing vs. releasing responses, button stiffness), CancelTime offers a more precise window into when inhibitory processes are enacted. Further, CancelTime has demonstrated good reliability, even with a lower number of trials (~15).^17^
